# Disease progression modeling in Alzheimer’s disease: insights from the shape of cognitive decline

**DOI:** 10.1101/2019.12.13.19014860

**Authors:** Lars Lau Raket, for the Alzheimer’s Disease Neuroimaging Initiative

## Abstract

**Background:** The characterizing symptom of Alzheimer disease (AD) is cognitive deterioration. While much recent work has focused on defining AD as a biological construct, most patients are still diagnosed, staged, and treated based on their cognitive symptoms. But the cognitive capability of a patient at any time throughout this deterioration will not directly reflect the disease state, but rather the effect of the cognitive decline on the patient’s predisease cognitive capability. Patients with high predisease cognitive capabilities tend to score better on cognitive tests relative to patients with low predisease cognitive capabilities at the same disease stage. Thus, a single assessment with a cognitive test is not adequate for determining the stage of an AD patient.

**Methods and Findings:** I developed a joint statistical model that explicitly modeled disease stage, baseline cognition, and the patients’ individual changes in cognitive ability as latent variables. The developed model takes the form of a nonlinear mixed-effects model. Maximum-likelihood estimation in this model induces a data-driven criterion for separating disease progression and baseline cognition. Applied to data from the Alzheimer’s Disease Neuroimaging Initiative, the model estimated a timeline of cognitive decline in AD that spans approximately 15 years from the earliest subjective cognitive deficits to severe AD dementia. It was demonstrated how direct modeling of latent factors that modify the observed data patterns provide a scaffold for understanding disease progression, biomarkers and treatment effects along the continuous time progression of disease.

**Conclusions:** The suggested framework enables direct interpretations of factors that modify cognitive decline. The results give new insights to the value of biomarkers for staging patients and suggest alternative explanations for previous findings related to accelerated cognitive decline among highly educated patients and patients on symptomatic treatments.

## 1. Background

Alzheimer disease (AD) is slowly progressing with preclinical and prodromal phases lasting many years before the onset of dementia. The stage of the underlying disease process of an AD patient entering a clinical trial is largely unknown, but may be estimated by a combination of, for example, cognitive testing, clinical evaluation, and biomarker results. While these procedures for evaluating disease severity are useful for creating coarse groupings of patients, the factors used to create groupings may be systematically affected by a wealth of factors not directly tied to the disease process, for example, intelligence, level of education, comorbidities, and genetics.

So far, efforts to develop therapies that delay or halt the progression of AD have generally been unsuccessful, and the vast majority of trials testing symptomatic agents in AD have also failed. These failures may be due to wrong therapeutic targets or non-efficacious therapies, but it is possible that a proportion of trial failures could be attributed to other factors such as study design, endpoints, and non-optimal patient population selection. For disease-modifying drugs, for example, the current standard durations for interventional studies may not be adequate. Simulations based on cohort studies suggest that prevention of disease in cognitively normal individuals may require study lengths far beyond the current standard to achieve high statistical power for detecting an effect of even very efficacious drugs [1,2], and this may also be the case for secondary prevention studies.

### 1.1 Cognitive decline and symptom onset

The characterizing symptom of AD is cognitive deterioration. The cognitive capability of a patient at any time throughout this deterioration will not directly reflect the disease state, but the effect of the cognitive decline on the patient’s predisease cognitive capability.

Many factors influence instantaneous cognitive ability, and low cognitive ability at a single time point is not necessarily an indication of cognitive decline. Cognitive decline can only be established by repeated evaluations of patients’ cognition over time. Longitudinal assessments of patient cognition also offer the benefit of hindsight – once cognitive decline is established; one can traverse back in time along the cognitive trajectory and predict when the decline started and search for patterns that are indicative of future cognitive decline. If done properly, one can synchronize individual observed trajectories to one long-term timeline representative of the full span of cognitive decline over the disease

Age is typically considered the major risk factor for developing AD, but age of first diagnosis of AD can vary by decades between patients, and because this span is much greater than the entire course of cognitive decline associated with AD, patient age is not an appropriate scale for understanding the pattern of cognitive decline in AD. The natural scale for studying the patterns of cognitive decline is time since symptom onset. However, self- or caregiver-reported age of symptom onset may be imprecise due to the patient’s memory problems; recall bias, where early sporadic cognitive issues are believed to be symptoms of the disease; or personal differences in sensitivity and interpretation of the earliest cognitive problems.

### 1.2. Disease progression modeling

Alzheimer disease typically presents in a sporadic late-onset form. The autosomal dominant forms of AD (ADAD) caused by rare genetic mutations have earlier onset than sporadic AD, but otherwise the pathogenesis is largely similar [3]. In ADAD, age at symptom onset is strongly affected by mutation type, parental age at symptom onset, APOE genotype and sex [4]. These factors can be used to calculate expected patient age at symptom onset for ADAD patients, which can be used to construct a more synchronized time scale for studying biomarkers and the pathological cascade of the disease [5]. Furthermore, this makes it possible to do primary prevention studies in a highly efficient manner [6].

In sporadic AD, age at onset cannot be predicted accurately from demographic or genetic factors. Assessment of biomarkers such as amyloid and tau load in cerebrospinal fluid or by positron emission tomography may be used to diagnose the disease even in the earliest stages [7], but such assessments can be both invasive and expensive, and data is sparse. There are however rich datasets with longitudinal cognitive measurements that span different parts of the disease. An appealing use of this data is to assemble the individual observed short-term trajectories to one long-term timeline representative of the full span of cognitive decline over the disease.

Different approaches to construct disease progression models for AD have been taken. A classic approach is to formulate the changes in cognitive scores using differential equations [8,9,10,11]. One major drawback of this type of modeling is that covariate effects and different sources of random variation should be formulated in the differential equation framework and may be very difficult to handle and interpret. An alternative class of disease progression models is based on direct modeling of the observed longitudinal trajectories and explicit modeling the patient-level disease stage. An important example of this type of approach is the model by Donohue et al. [12] which simultaneously models multiple observations of cognitive measures and biomarkers. This modeling approach has been powerful in illuminating the multivariate nature of Alzheimer disease progression, but it does have some drawbacks. In particular, the model assumes that all included outcomes are synchronized over a common disease time scale; it does not model correlations between outcomes; it does not include covariates; and finally, it uses a heuristic estimation procedure that does not simultaneously account for the assumed random variability in both timing and measurements. The approach was recently generalized to a wider class of Bayesian latent-time joint mixed-effects models [13]. This class of models can in principle take advantage of the dependence between different outcomes and allow inclusion of covariates, but covariates can only model variation in outcomes and not disease stage or progression rate. Furthermore, the Bayesian element of the model requires somewhat arbitrary choices of priors for different model parameters. Progression is modeled on an age scale and, as discussed above, this choice may amplify the a-priori dis-synchronization between patients by orders of magnitude. This in turn negatively affects the results of the model as can be seen in Figure 1 in the paper [13] where patient-level trajectories go from minimal to maximal severity over 10-15 years, while variation of when maximal severity is reached between patients is spread out over 20-year periods.

**Figure 1.**
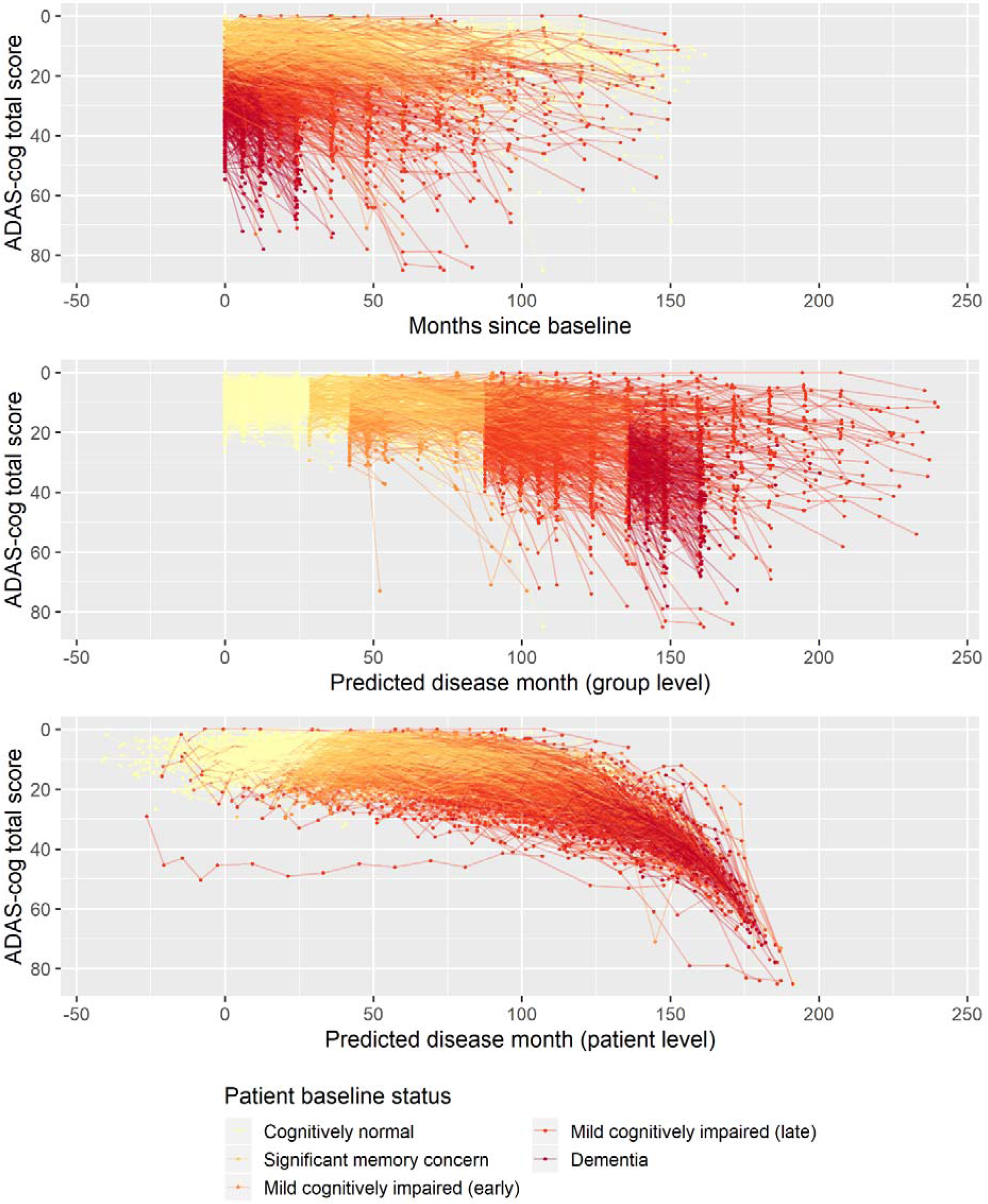
Observed longitudinal ADAS-cog trajectories for 2,142 ADNI participants plotted against time in study (top), predicted patient baseline-status group-level disease time (middle), and predicted individual disease time.

In this paper, we propose a new approach to disease progression modeling that overcomes several of the shortcomings of the above-mentioned methods. In the presented form, the model is estimating a disease timeline from repeated assessments of a univariate measure such as a cognitive scale. The model is inspired by the statistical framework presented by Raket et al. [14] where systematic patterns of variation in both vertical (observed cognitive score) and horizontal (disease-timing) directions are modeled simultaneously on both the population and individual level. The model allows covariate effects on both outcomes and disease progression and all model parameters are estimated simultaneously using maximum likelihood estimation.

The goal of this work was to explore whether the proposed disease progression model could align observed cognitive trajectories to a precise timeline of cognitive decline associated with AD and to evaluate if this modeling would shed new light on aspects related to disease progression and biomarkers. When the model was fitted to cognitive scores from ADNI, the presented model aligned the cognitive trajectories of patients to a consistent shape of cognitive decline with a span of approximately 15 years from the earliest subjective cognitive deficits to severe AD dementia. It was shown that the model’s predictions of patients’ disease stages based on their longitudinal cognitive scores could predict time since symptom onset and diagnosis. It was further demonstrated that the predicted disease stages provided a more suitable time scale for modeling the evolution of biomarkers over the course of disease than group-wise modeling based on patient symptoms at baseline. The model was used to estimate the effects of sex, age, and education on cognitive decline and to evaluate the effects of cholinesterase inhibitor treatment on cognitive decline. Finally, the model was fitted to the cognitive trajectories of a subset of patients with a rich set of biomarkers available at baseline to estimate if baseline biomarker profile could predict disease stage. The results of the model in an independent held-out validation data set confirmed that baseline biomarker profiles could predict the disease stage of unseen individuals – even in the preclinical phases of disease where no clinically detectable cognitive impairment is present.

## 2. Methods

### 2.1. Data

Data used in the preparation of this article were obtained from the Alzheimer’s Disease Neuroimaging Initiative (ADNI) database (adni.loni.usc.edu). The ADNI was launched in 2003 as a public-private partnership, led by Principal Investigator Michael W. Weiner, MD. The primary goal of ADNI has been to test whether serial magnetic resonance imaging (MRI), positron emission tomography (PET), other biological markers, and clinical and neuropsychological assessment can be combined to measure the progression of mild cognitive impairment (MCI) and early Alzheimer’s disease (AD). For up-to-date information, see www.adni-info.org.

Patients included in the current study were required to have a valid classification at baseline (cognitively normal, significant memory concern, mild cognitive impairment [early], mild cognitive impairment [late], or dementia).

#### Outcomes

The main outcome measure considered was the total score of the 13-item Alzheimer’s Disease Assessment Scale-cognitive subscale (ADAS-cog; range: 0-85; lower score indicates less impairment) [15]. Included patients were required to have at least one valid ADAS-cog total score to be included in the present study.

Other outcomes were reported onset of various symptoms related to cognitive impairment and AD; Clinical Dementia Rating scale – sum of boxes [16]; Functional Activities Questionnaire [17], FDG-PET (meta-ROI) [18]; cross-sectional hippocampal volume extracted from MRI using FreeSurfer [19]; Florbetapir PET SUVr [20]; Aβ_1-42_, total tau, and p-tau_181_ concentrations in CSF as measured using the Roche Elecsys^®^ immunoassay [21]; the ratio of Aβ_1-42_/Aβ_1-40_ concentrations in CSF as measured by 2D-UPLC tandem mass spectrometry; and Neurofilament Light chain (NfL) levels in plasma measured using a single molecule array platform [22].

### 2.2. Disease progression model

Let *y*_*ij*_ represent the observed cognitive score of patient *i* at time *t*_*ij*_(*i* = 1, …, *n, j* = 1, …, *m*_*i*_). We assume that *y*_*ij*_ is generated by a model of the form

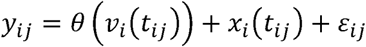

where *θ* is a function that represents the shape of cognitive decline; *v*_*i*_ is a warping function that transforms observation time *t*_*ij*_ to a disease time scale *v*_*i*_(*t*_*ij*_) that is aligned across patients; *x*_*i*_ is the idiosyncratic patient-level deviation from the mean shape that represents consistent deviations over time; and *ε*_*ij*_ is independent measurement noise.

Cognitive scores can be extremely noisy due to many different sources of variation, and to accurately infer the shape of the disease timeline of cognitive decline *θ*, to predict patient-level disease stage, and to predict the entire patient-level course of decline *ŷ*_*i*_, one will have to make suitable model choices. In the following, we describe the basic model choices and their motivations.

Because we are modeling cognitive decline in a pathological aging, it is natural to assume that the representative shape of decline *θ* is a function that has a stable left asymptote (predisease cognitive normality) and a monotone decline. In this paper, we focus on ADAS-cog scores that show a distinct exponential decline in dementia [23], and thus we will work with a parametrized family of exponential functions to model the mean progression pattern

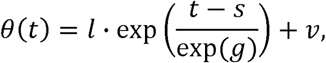

where *v* is the left asymptote representing the average stable predisease cognitive score, and where the remaining parameters determine the shape of the decline.

The mean progression pattern *θ* can be modeled differently to achieve other properties, for example as a generalized logistic function or as a monotone spline [24].

The mapping of observed time to disease time *v*_*i*_ should allow the model to assemble short-term longitudinal observations to a long-term timeline of cognitive decline. The major source of variation can likely be ascribed to differences in how long the patient has had the disease before we begin observing them, so we model *v*_*i*_ as a shift of study time

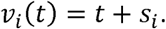

#### Random effects

When modeling longitudinal data for groups of individuals it is often natural to describe systematic differences between individuals using random effect. The proposed disease progression model has three types of random effects.

- *s*_*i*_: random patient-level shift that models the disease stage of patient *i*. Assumed to follow a zero-mean normal distribution with unknown variance.
- *x*_*i*_: random patient-level systematic deviation from the mean curve. Assumed to be a discrete-time observation of a Brownian motion with a zero-mean normally distributed starting level with unknown variance. The Brownian motion has an unknown parameter controlling variance scale.
- *ε*_*ij*_: random observation noise. Assumed to be independent zero-mean normally distributed with unknown variance.

A free correlation between *s*_*i*_ and the normally distributed starting level of *x*_*i*_ is included in the model, the remaining effects are assumed independent.

#### Fixed effects

The basic model parameters *l, g, s*, and *v* that describe the shape of *θ* are modeled as fixed effects.

- *l* is a scaling parameter of the exponential function. Since a goal of disease progression modeling is to find a common pattern of decline, *l* will be modeled as a single free parameter.
- *g* is a scaling parameter of time. Patient-level differences in rate of decline that can be ascribed to a covariate or factor can be modeled as a regression-type model on *g*. Initially this parameter will be modeled as a single free parameter.
- *s* is a shift of observed time. Patient-level differences in disease stage that can be ascribed to a covariate or factor can be modeled as fixed effects. Since the present study includes several cohorts at different disease stages (e.g. cognitively normal patients, patients with dementia) the initial modeling will have different *s* parameters for non-cognitively normal cohorts, thus modeling disease time since the baseline stage of the cognitively normal patients.
- *v* is an intercept parameter describing the left asymptote. Patient-level differences in predisease cognition that can be ascribed to a covariate or factor can be modeled as a regression-type model on *v*. Initially this parameter will be modeled as a single free parameter.

### 2.3. Statistical analysis

To investigate the effect of covariates on the pattern of disease progression, forward selection was used to evaluate models with all combinations of covariate effects on rate of decline *g*, disease stage *s*, and predisease cognition *v*. The search was continued as long as the Akaike Information Criterion [25] improved, but the model selection was based on the more conservative Schwarz’ Bayesian Information Criterion (BIC) [26].

To investigate if predicted disease time was predictive of time since reported symptom onset, linear regression was done on time since reported symptom onset (at baseline) using predicted disease time as a covariate. P-values were computed using T-tests.

To investigate if predicted disease time offered a better time scale for modeling other longitudinal outcomes (e.g. biomarkers) than time since baseline for the five baseline groups, linear mixed effects modeling was used. To allow for nonlinear trends in the mean pattern, the outcome was modeled using a cubic B-spline function with 3 degrees of freedom across predicted disease time and time since baseline (one pattern per baseline group), respectively. Patient-level random slopes and intercepts were included to model longitudinal deviations within an individual. P-values were computed using likelihood ratio tests with maximum likelihood estimation.

Comparisons of quantitative outcomes between groups with two levels were done using Wilcoxon rank sum tests and correlations were evaluated with Spearman’s rank correlation coefficients.

#### Software

All analyses were done using R version 3.5.2 [27]. Maximum likelihood estimation in the disease progression models was done using the method of Lindstrom and Bates [28] using the nlme and covBM R packages [29].

## 3. Results

### 3.1. Basic model

The basic model described in Section 2.2 was fitted on longitudinal ADAS-cog data from ADNI. The data comprised 9,830 ADAS-cog scores across 2,142 individuals. The ADAS-cog scores plotted against study time and predicted time-scales (“disease month”) are shown in Figure 1. Relative to the average baseline disease stage of the Cognitively normal group the model estimated that the Significant memory concern group were 29 months later into the trajectory of cognitive decline, while the early and late MCI groups were respectively 42 and 88 months later, and that the dementia group was 136 months later. The model had 12 degrees of freedom, and the log likelihood of the fitted model was -29734.26. AIC and BIC were 59492.52 and 59578.84 respectively.

### 3.2 Validation of the basic model

The presented disease progression model aggregates the information in baseline status groups and the longitudinal trajectories of participants to a single number, the predicted disease month. For this continuous disease progression scale to relevant to AD, it should also hold information that describe other aspects of the disease than the cognitive deterioration observed on ADAS-cog that the model was fitted on.

To first evaluate whether the disease progression model captured milestones of cognitive deterioration, we evaluated the model’s ability to predict self-reported onset of cognitive symptoms, mild cognitive impairment symptoms, Alzheimer’s disease symptoms or diagnosis of Alzheimer’s disease. There were 1.142 participants who had at least one entry of these data during the study follow-up. Age of symptom onset or diagnosis plotted against the age at the model’s predicted disease time 0 is shown in Figure 2. Ideally (based on perfectly aligned trajectories and perfect self-reported onsets/diagnosis between individuals) the results of each measure would lie on a line with slope 1 and the intercept would represent the difference in years between age at disease time 0 and the age at onset/diagnosis time. For the age of onset of cognitive symptoms, there seems to be different intercepts for the different baseline groups, where more severe baseline groups tend to report symptom onset later relative to the model prediction of the less severe groups. This may be an effect of different subjective definitions of onset of cognitive symptoms across baseline groups; it may be because of biased model estimates of the staging of the baseline groups; or a combination.

**Figure 2.**
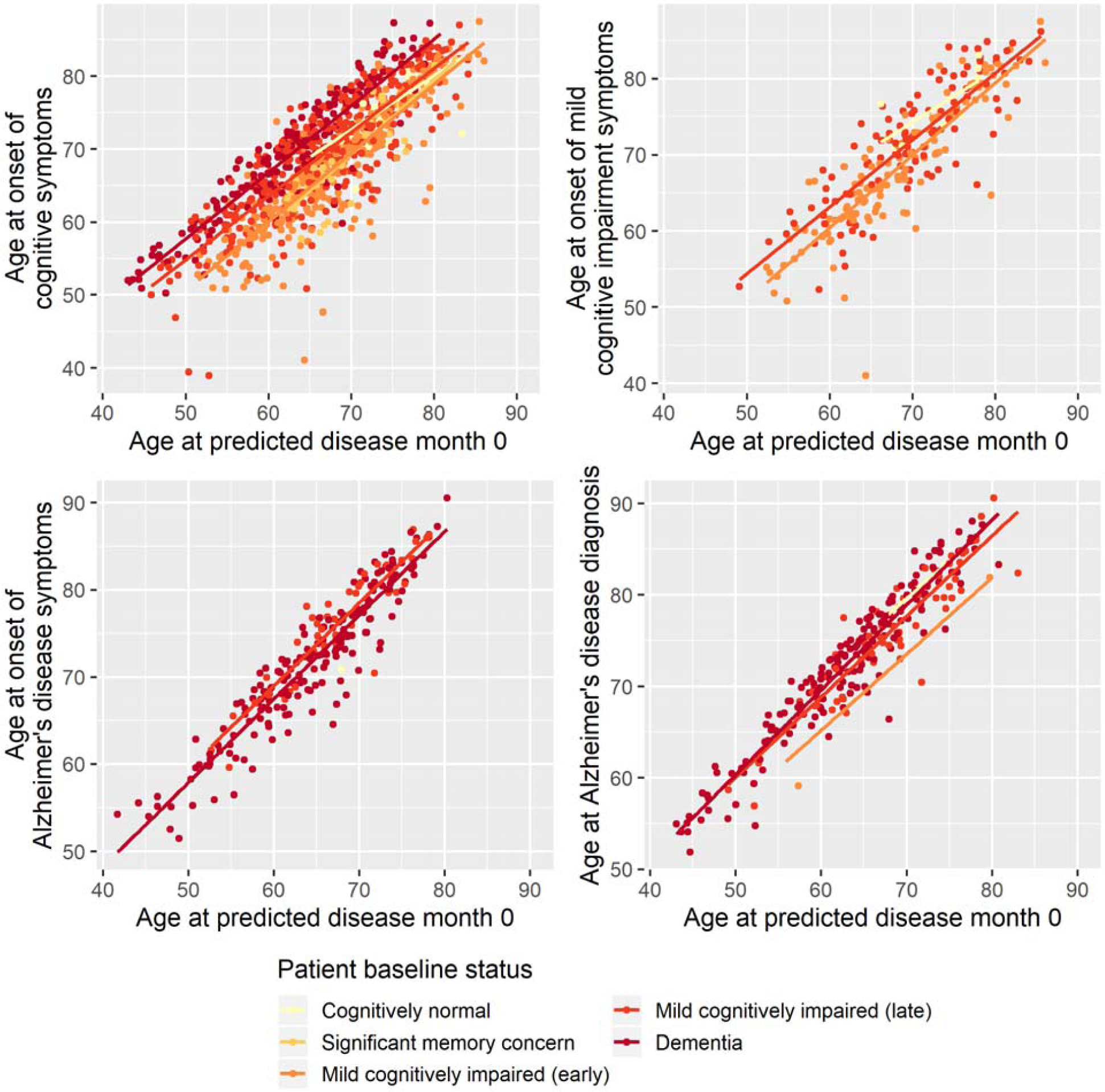
Reported age of onset of cognitive symptoms (top left), cognitive impairment symptoms (top right), Alzheimer’s disease symptoms (bottom left) and Alzheimer’s disease diagnosis (bottom right) as a function of age at predicted disease month 0.

Based on linear regression, predicted disease month was predictive of time since cognitive symptom onset (p < 0.0001), time since Alzheimer’s disease symptoms onset (p < 0.0001) and time since Alzheimer’s diagnosis (p < 0.0001) – all times relative to study baseline. Predicted disease month was not significantly predictive for time since mild cognitive impairment symptom onset (p = 0.558).

Secondly, to validate that the predicted disease time also synchronizes other independently captured aspects of the disease than cognition as measured by ADAS-cog, we analyzed if this continuous disease scale based on baseline patient statuses and ADAS-cog trajectories better captured patterns of variation in other clinical scales and biomarkers than separate modeling of the different baseline groups. We found that the predicted disease time better described the patterns of variation compared to allowing separate patterns per baseline group in 7 of the 10 cases when measured by log likelihood (Table 1), even though the latter model had 16 degrees of freedom more than the former. When measured using AIC and BIC that both adjust for additional degrees of freedom to compare model quality, the predicted disease time model was better in 8 of the 10 cases for AIC and 10 of the 10 cases for BIC. Interestingly, the three biomarkers where group-wise modeling was better as measured by log likelihood were all measuring to beta amyloid (CSF Aβ_1-42_ and Aβ_1-42_/Aβ_1-40_ ratio, Florbetapir PET). These biomarkers are known to have a bimodal distribution [30] and are thus poorly modeled by a single trajectory. Figure 3 shows the estimated trajectories of the two models for hippocampal volume (MRI), Aβ_1-42_ (CSF), and NfL (plasma).

**Table 1.**
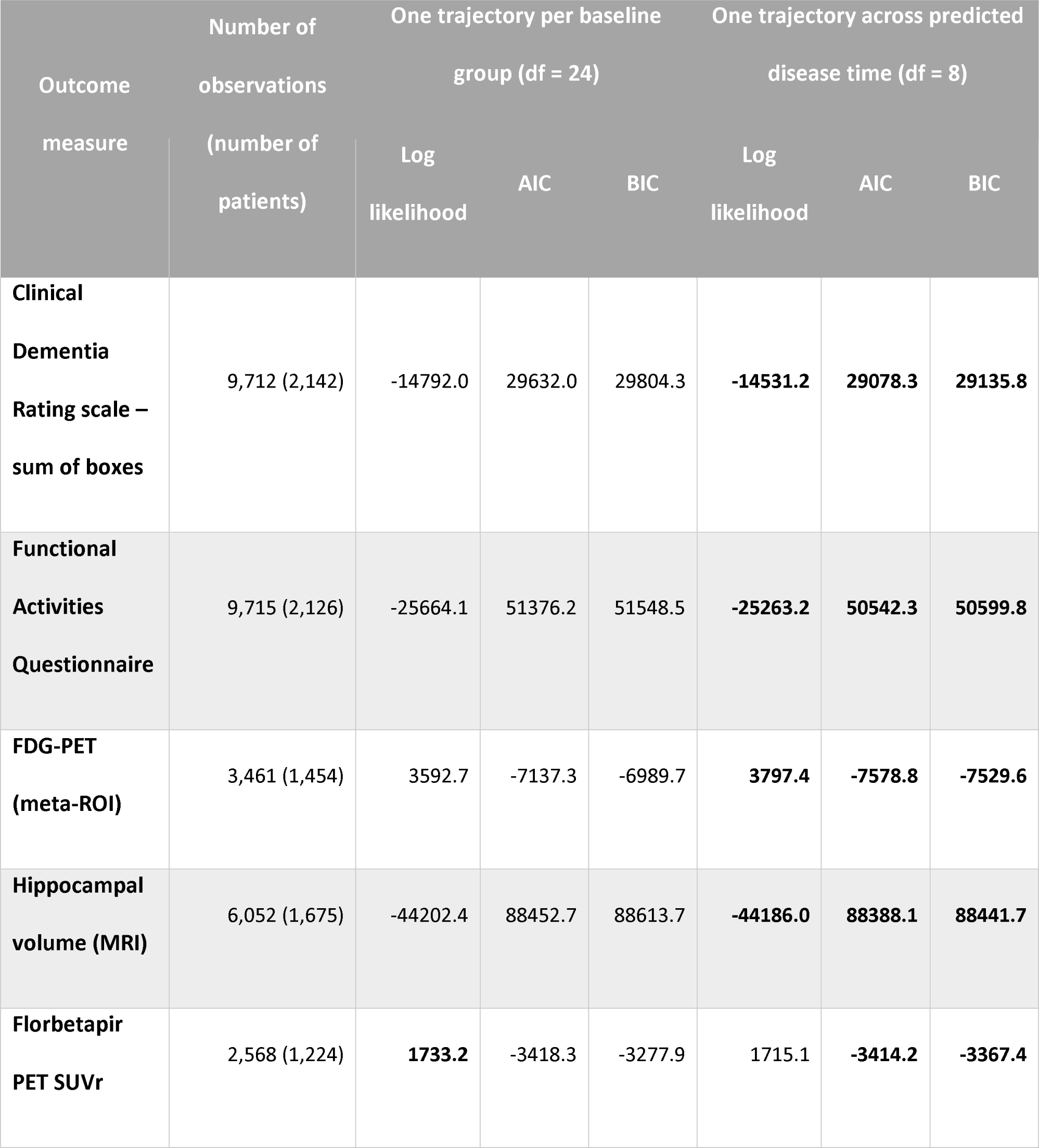
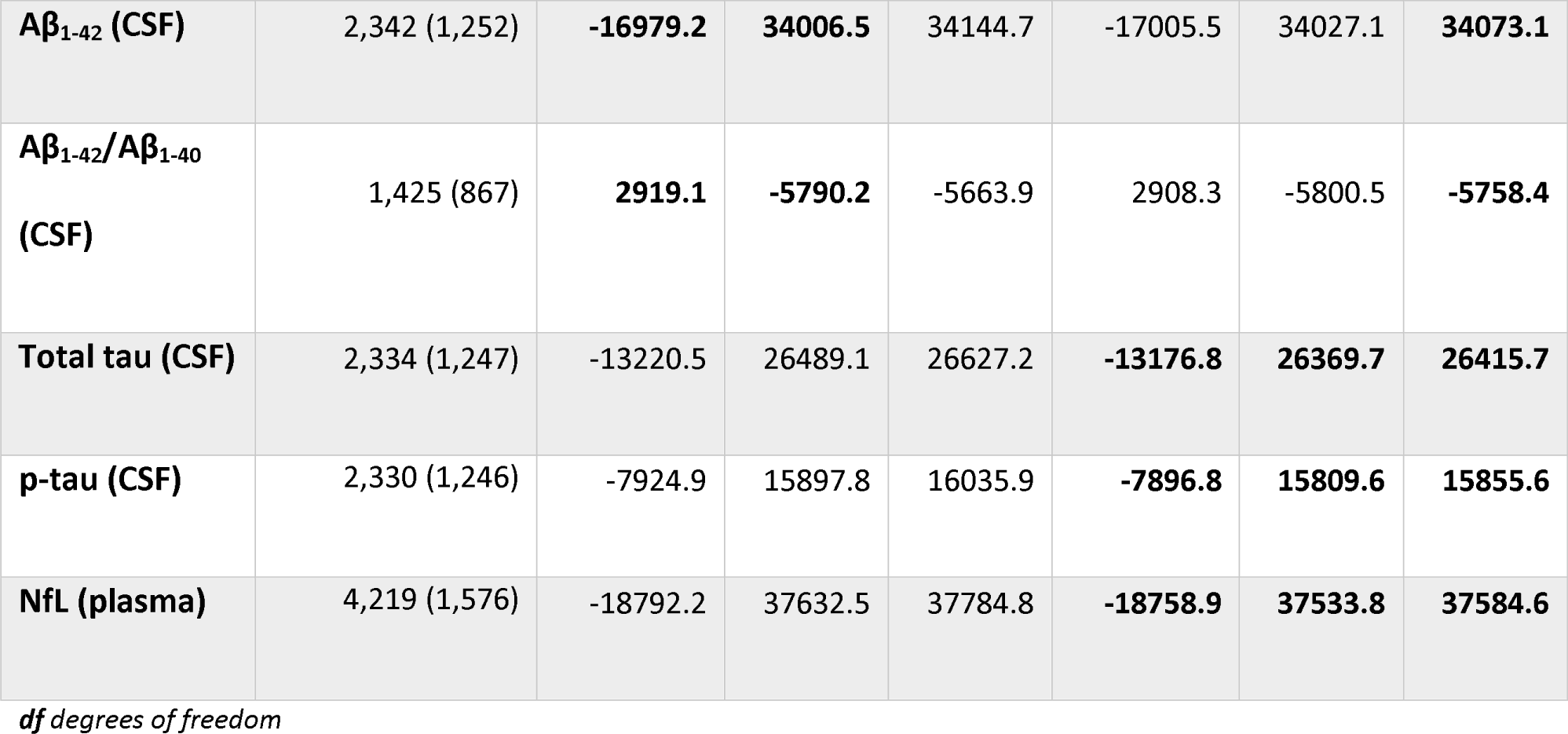
Comparison of longitudinal modeling of clinical scales and biomarkers based on patient baseline group versus continuous disease time. Comparison in terms of log likelihood (larger is better), AIC and BIC (smaller is better). Bold numbers indicate the best fitting model for a given measure.

**Figure 3.**
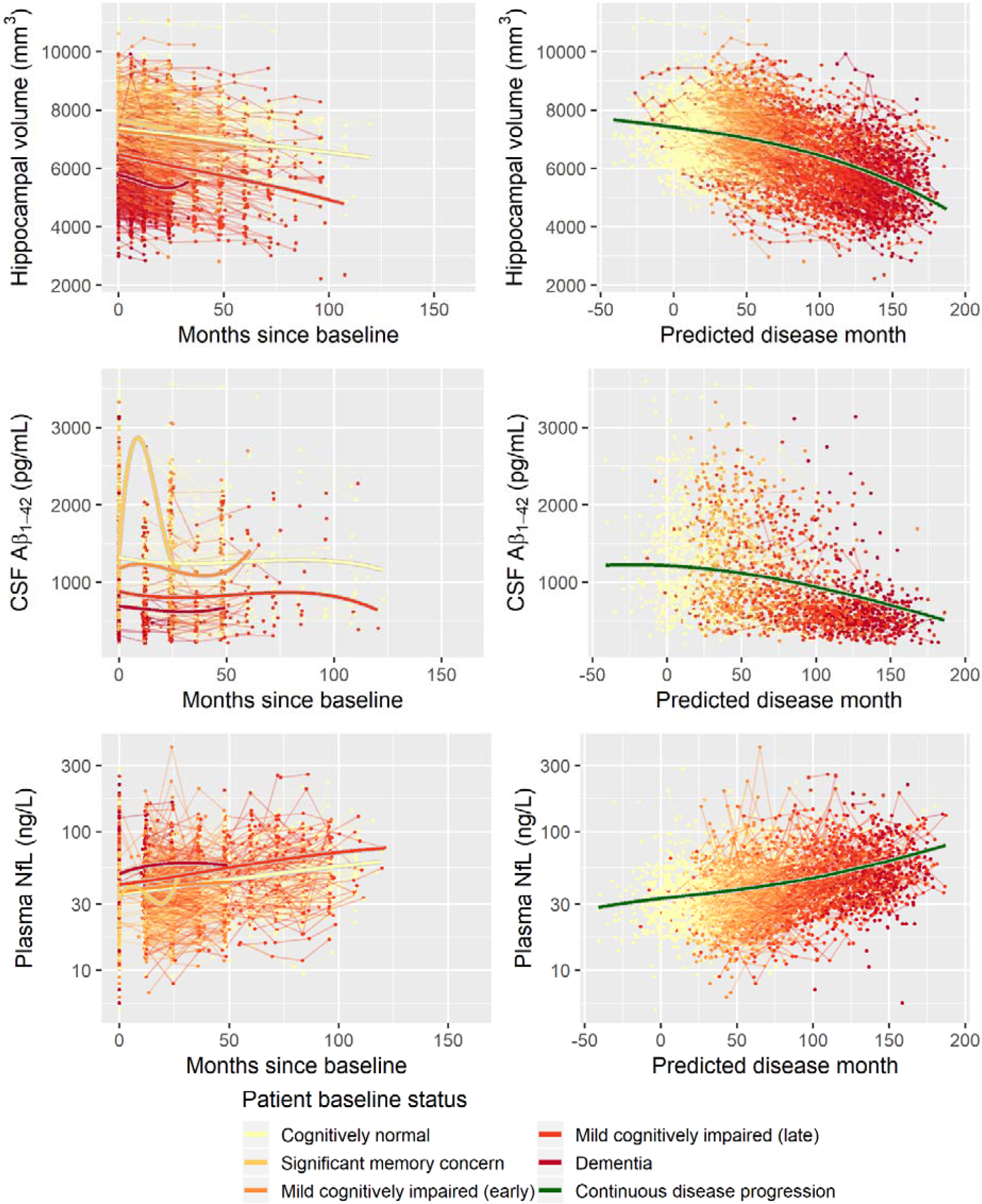
Data and estimated biomarker trajectories for hippocampal volume (MRI), Aβ_1-42_ (CSF), and NfL (plasma). Left column shows results when allowing different trajectories for the five baseline groups, right column shows the results when requiring a single trajectory over predicted disease time.

### 3.2 Age, sex, education and cognitive decline

There were systematic differences in follow-up time, age at baseline and length of education between male and female participants (**Supplementary Table 1**). Compared to female participants, male participants on average had 3.2 months longer follow-up (Wilcoxon p = 0.0085), were on average 2.0 years older at baseline (Wilcoxon p < 0.0001) and had 0.89 years more education (Wilcoxon p < 0.0001). Age at baseline and years of education were not significantly correlated (Spearman’s ρ = –0.04, p = 0.0792).

To explore whether age at baseline, sex and length of education affected the pattern of cognitive decline, stepwise forward model selection was done to include these factors in the model. The best model included fixed effects of age and sex on *g,s* and *v*, and fixed effects of years of education on *g* and *v*. While there were some substantial differences in marginal parameter estimates due to age, sex and length of education (e.g. men are predicted to be 57 months later in disease compared to women in the same baseline groups; **Supplementary Table 2**), the estimates should not be interpreted in isolation since all parameters simultaneously affect the shape of the disease trajectory and may counteract each other. Figure 4 shows how age, sex and education differences systematically affected the mean trajectories. From the figure we see that male participants consistently scored lower on ADAS-cog throughout the disease (3.1 points), but that they remained more stable in the initial 100 months where female participants had a more gradual decline. Lower age at baseline and longer education were both associated with higher cognitive scores, but also slightly increased rates of decline as evident in the stages of overt dementia (predicted disease time >120 months).

**Figure 4.**
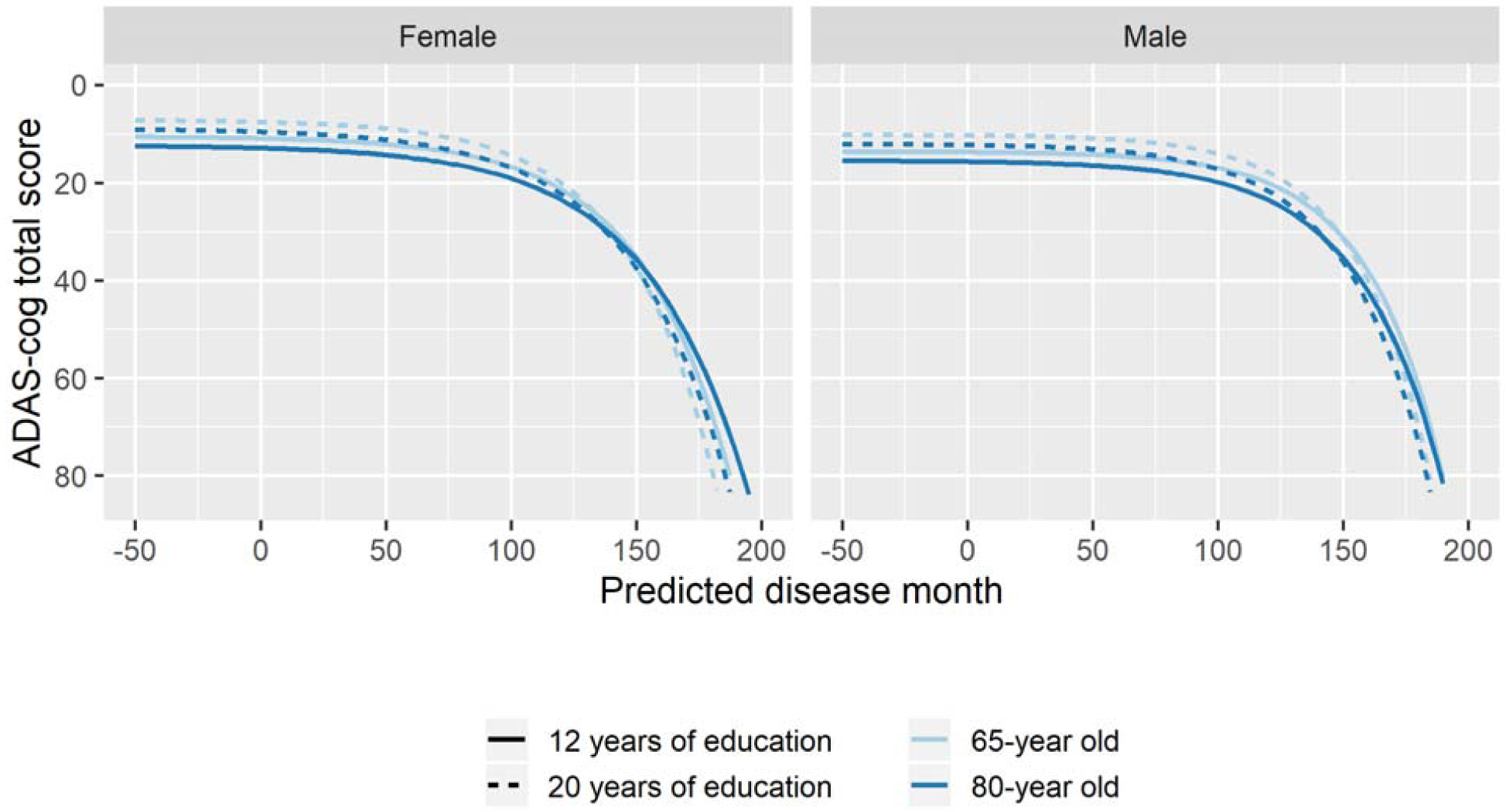
Estimated trajectories for different combination of patient age, sex and length of education. The trajectories are aligned at predicted month 0 that corresponds to the average cognitive stage of cognitively normal individuals at baseline.

### 3.3 Cholinesterase inhibitors and cognitive decline

Using the search terms described in the supplementary material, we identified 1,347 individuals that were treated with cholinesterase inhibitors (ChEIs), but only 64 of these initiated or discontinued treatment during the observation time (total of 9 initiations and 60 discontinuations).

To explore if treatment with ChEIs affected the shape of the decline trajectories, stepwise forward model selection from the basic model was done to include ChEI treatment in the model. The best model included fixed effects of treatment on *s* and *v*, but not on rate of decline *g* (14 degrees of freedom, log likelihood = -29638.79, AIC = 59305.59, BIC = 59406.29). The model found that patients treated with ChEIs generally had worse level of cognition (effect on *v* was 5.50 ADAS-cog points for treated individuals, p < 0.0001) and a delayed progression within baseline groups (effect on *s* was 7.53 months, p < 0.0001). The average trajectories and distribution of data across treatment are shown in Figure 5.

**Figure 5.**
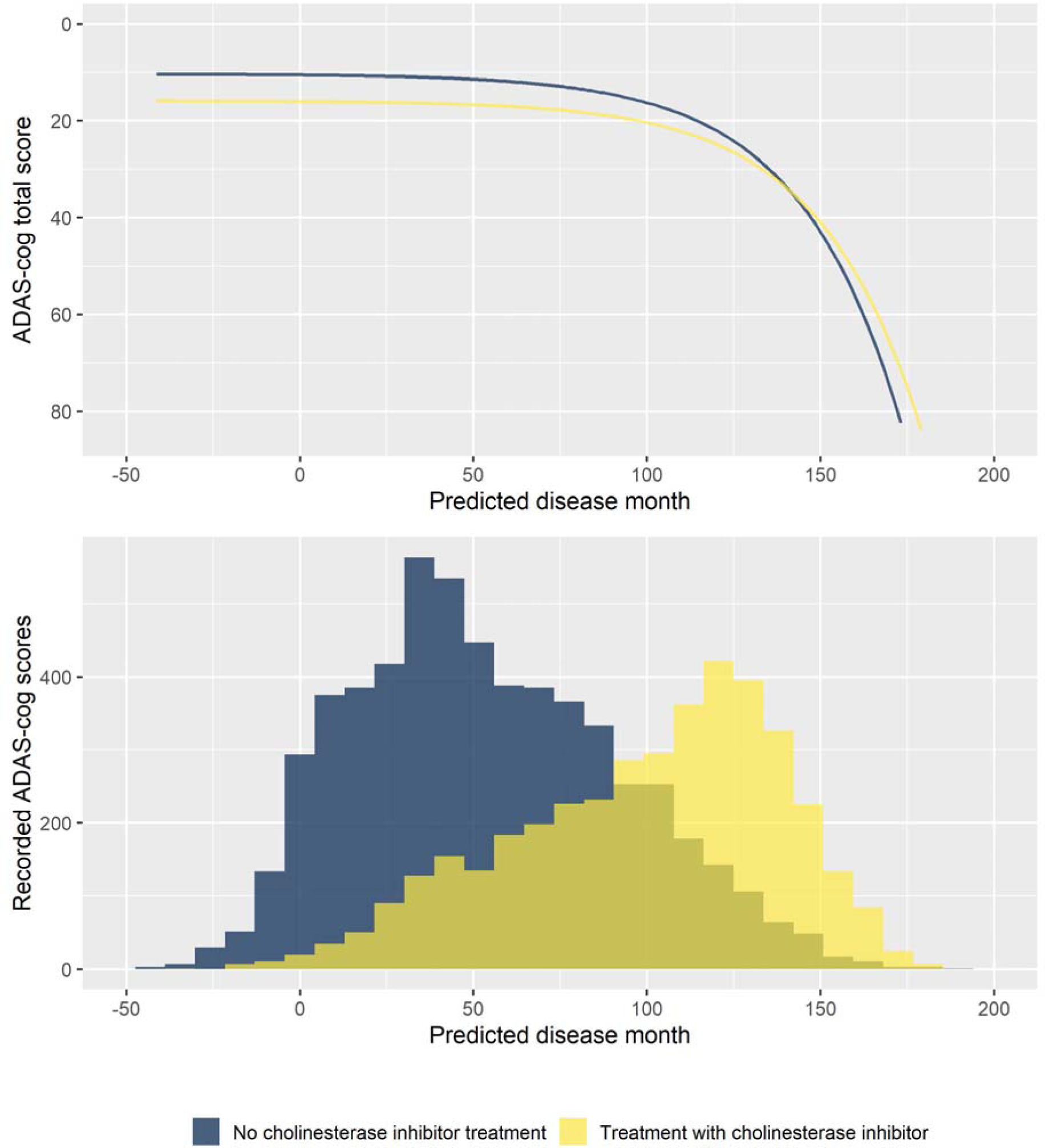
Estimated trajectories for patients with and without cholinesterase inhibitor treatment (top) and corresponding distribution of observed ADAS-cog scores. The trajectories are aligned at predicted month 0 that corresponds to the average cognitive stage of cognitively normal individuals that are not treated with cholinesterase inhibitors at baseline.

### 3.4 Biomarkers for disease staging

The disease progression model relies on observing patients longitudinally and uses the temporal pattern of cognitive scores to predict the patient’s status at baseline. This is valuable for increasing disease understanding for cohorts or for doing retrospective analyses, but the models presented thus far offer only little insight into the disease stage of a patient entering, for example, a clinical trial. In this setting, only the baseline classification of the patient, the cognitive score and possibly other demographic data would be able to inform the stage of the patient. However, as shown in Section 3.2, several biomarkers have temporal patterns that follow the trajectory of cognitive decline. Biomarker data collected at baseline may thus enable a better assessment of the stage of an individual.

688 individuals had complete biomarker data at baseline for the eight biomarkers considered in Section 3.2. These individuals had 3,301 visits with valid ADAS-cog scores.

#### Training and validation data

540 individuals (80%) were randomly selected for the training cohort and the remaining 148 (20%) comprised the validation cohort.

#### Model development

Using the BIC-based model selection procedure described previously, we searched for the best model among models that included adjustment for sex, baseline age and education (on parameters *g, s, v*) as well as adjustment for the eight baseline biomarkers on disease stage (parameter *s*). The model selection was done on the training data. The best model included the biomarkers FDG-PET (meta-ROI), hippocampal volume (MRI), Florbetapir PET SUVr, Aβ1-42/Aβ1-40 (CSF), and NfL (plasma) (22 degrees of freedom, log likelihood = -7591.17, AIC = 15226.34, BIC = 15355.45). The parameter estimates for the model are given in Supplementary Table 3.

#### Model validation

To validate the biomarker model, the model fitted on training data was used to predict disease stage in two different scenarios. The first used baseline biomarker data and the second used baseline biomarkers in combination with baseline ADAS-cog total score. Visual inspection of the longitudinal ADAS-cog trajectories suggests that the baseline data does hold information that improves prediction of disease stage in the test data (Figure 6). To quantify this, the biomarker model was compared to the baseline model on its predictive accuracy for the longitudinal ADAS-cog total score trajectories (Table 2). Inclusion of biomarker data clearly reduced the mean squared error (MSE) and median absolute error (MAE) of predictions on both test and training data (MSE/MAE 65.1/4.21 vs. 100.0/4.98 on test data). Including the baseline ADAS-cog total score improved the post-baseline predictive accuracy of the biomarker model further (MSE/MAE 55.1/3.48 for baseline biomarkers + ADAS-cog model vs. 69.8/4.31 for biomarker model on test data).

**Table 2.** Predictive accuracies of predicted ADAS-cog total score trajectories for the basic model and the biomarker model both with and without the baseline ADAS-cog total score. Predictions were censored to the interval [0, 85] to respect the range of the ADAS-cog scores.

| Model and data | Mean squared error |  | Median absolute error |  |
| --- | --- | --- | --- | --- |
|  | Training | Test | Training | Test |
| **Basic model**<br>**(baseline status)** | 90.0 | 100.0 (110.6<sup>1</sup>) | 5.48 | 4.98 (5.01<sup>1</sup>) |
| **BM model**<br>**(baseline status +**<br>**BM)** | 58.3 | 65.1 (69.8<sup>1</sup>) | 4.19 | 4.21 (4.31<sup>1</sup>) |
| **Basic model**<br>**(baseline status +**<br>**ADAS-cog)** | 78.2<sup>1</sup> | 102.7<sup>1</sup> | 3.49<sup>1</sup> | 3.70<sup>1</sup> |
| **BM model**<br>**(baseline status +**<br>**ADAS-cog + BM)** | 53.5<sup>1</sup> | 55.1<sup>1</sup> | 3.29<sup>1</sup> | 3.48<sup>1</sup> |

*BM: Biomarker*
<sup>1</sup>Baseline ADAS-cog measurements excluded in computation of prediction errors

**Figure 6.**
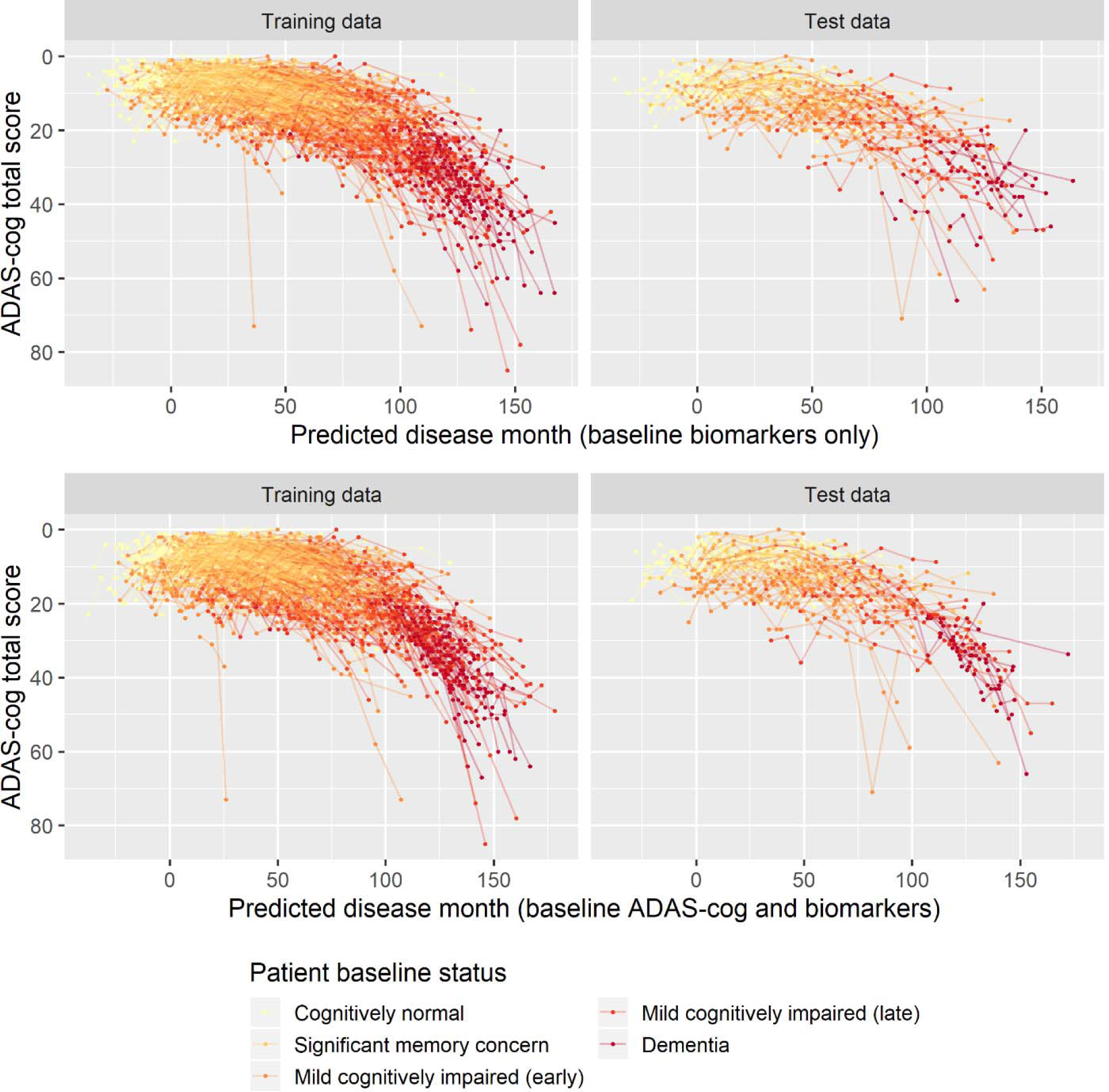
Predicted disease month for training and test datasets. Top row displays predicted disease-time alignment of observed ADAS-cog total score trajectories based on baseline biomarker data; bottom row displays predicted disease-time alignment of trajectories based on baseline ADAS-cog total score and baseline biomarker data

## 4. Discussion

### Disease progression modeling

In this paper we presented a model for progression of dementia based on longitudinal cognitive assessments. Disease stages of individual patients were modeled using a latent variable approach. As opposed to conventional latent variable models, for example, in item response theory for modeling cognitive tests [31,32], the proposed model imposes explicit structures to ensure that the longitudinal modeling respects the known course of disease (e.g. that disease progression is an increasing function of elapsed time and that cognition on average declines with disease progression). By imposing these structures, the model provides a scaffold for understanding disease progression in pathological aging in terms of three continuous measures, *disease stage, rate of decline, and cognitive deviation from the mean*.

The proposed model aligned trajectories of cognitive decline, the characterizing symptom of dementia, and to demonstrate that this approach provided valid insights about other aspects of the disease, it was shown that predicted disease stage was predictive of various measures of disease onset. Furthermore, the use of ADAS-cog trajectories to map patients to a one-dimensional disease timeline consistently provided a better explanation of other clinical scales and biomarker trajectories than a conventional approach that grouped patients based on baseline symptoms.

### Age, sex, education and cognitive decline

The effect of demographic and socioeconomic factors on disease risk and manifestation in Alzheimer’s disease has been the subject of much study. In this work we focused on the combined effects of age, sex and length of education.

When considered individually, these factors have been observed to result in differences in disease progression. While age is typically considered the major risk factor for developing AD, higher age at AD onset has been observed to be associated with a slower rate of cognitive decline [33,34]. Similarly, female sex has been identified as a major risk factor with almost two-thirds of AD cases being women [35]. While this difference has been known for a long time, it has only become apparent more recently that there are sex differences in symptomatology, rate of decline, and possibly in neural anatomy [36,37]. Finally, the effects of cognitive reserve on age-related cognitive decline has been the subject of much study [38]. Cognitive reserve is often studied using educational attainment as an operational proxy for cognitive reserve. It has consistently been found that higher education is associated with increased rate of cognitive decline in incident Alzheimer’s disease [39,40,41,42,43,44,45]. Several of these studies also report that education is associated with higher baseline cognition.

Differences in cognitive decline is often studied by comparing slopes in statistical models that assume that cognitive decline follows a linear pattern. The argumentation and interpretation around the cognitive reserve model is somewhat more sophisticated, but still largely centered on an assumption of a linear rate of decline (e.g. illustrated in Figure 1 in [46]). The prevailing hypothesis within the field of cognitive reserve research is that, compared to individuals with low cognitive reserve, individuals with high cognitive reserve have higher predisease cognitive scores and that their brains tolerate a higher load of neuropathology before cognitive decline is seen. At a sufficiently high level of neuropathology, cognitive ability reaches its floor for all participants. If the timescale of neuropathological buildup is similar across individuals, this suggests that individuals with high cognitive reserve will have to decline a wider range of cognitive scores in a shorter time, thus leading to an accelerated rate of decline [46].

The analyses in the present paper clearly illustrate that rate of cognitive decline as measured on ADAS-cog is not constant but increases over the course of AD. Thus, findings of an increased rate of decline in a certain group of patients using slope models could either be because the group of patients has accelerated decline, because they are at a later disease stage, or a combination. The proposed disease progression model seeks to align cognitive trajectories on a disease timeline, and thus it allows one to separate the hypothesized mechanisms of cognitive decline. The best model that adjusted for effects of age at baseline, sex, and length of education on, respectively, disease stage, rate of decline and cognitive deviation, found that that all three factors affected all three disease measures except for disease stage which was not affected by length of education.

When considering the combination of effects (Figure 4), the results suggested that higher age at baseline was associated with lower cognition throughout disease time and a slightly reduced rate of decline. Women tended to have better predisease cognition but also an accelerated decline. Finally, longer education was associated with slightly faster rate of decline and a systematically better cognition throughout the disease.

While these findings are largely consistent with previous findings, they also illustrate that previous results that do not take the long-term disease trajectories into account may be systematically biased. In particular, the fact that highly educated patients tend to have above-mean cognition throughout the disease means that they will meet cognitive cut-offs used for inclusion criteria in clinical studies longer in to their disease than patients with less education. Because of the accelerated cognitive decline in the later stages of disease, these patients will have a much faster rate of decline when using conventional slope models, but this difference will primarily be due to their later disease stage.

### Symptomatic medications for Alzheimer disease and cognitive decline

ChEIs have consistently shown a symptomatic benefit in mild to severe dementia due to AD in randomized, double blind, placebo-controlled trials [47]. It has however been questioned whether long-term treatment with ChEIs could be harmful [48]. A recent meta-analysis found that AD patients treated with symptomatic treatments had a faster rate of cognitive decline [49]. This could be interpreted as a harmful side effect, but since the included studies were not randomized with respect to symptomatic treatments, such causal link cannot be made. An alternative explanation is simply that ChEIs work – that patients that are being treated at study inclusion have a cognitive benefit that, similarly to higher levels of education, means that they meet inclusion criteria for clinical studies further in to their disease. The optimal disease progression model identified in the model search did not include effects of ChEI treatment on rate of decline. Instead, the results of this model showed that patients treated generally had lower cognition compared to untreated patients (confounding by indication) and that their progression was slightly delayed.

### Biomarker-based disease staging

The final application of the model examined how a patient’s biomarker profile at study entry could be used to predict their disease stage. Based on training data used for model development a set of 5 biomarkers were included in the model. Inclusion of biomarker profiles considerably improved prediction of future ADAS-cog trajectories in the unseen validation dataset and inclusion of baseline ADAS-cog score further improved the prediction. Among the biomarkers, FDG-PET explained most variation followed by CSF Aβ1-42/Aβ1-40 and Florbetapir SUVr. Hippocampal volume and plasma NfL explained the least.

This modeling of baseline biomarkers for patients in the earliest stages of disease takes advantage of the long-term follow-up that is unique to ADNI. The modeling essentially relies on hindsight because the patients’ disease stage can only be predicted with high reliability once a systematic pattern of cognitive decline has been observed. By using these patterns, the model identified how combinations of biomarkers could be used to predict disease stage. The results of the model suggest that biomarker profiles at a single time point may be used to predict the disease stage of an individual even in the preclinical phases of disease where no clinically detectable cognitive impairment is present.

With further validation, these results can be used to define a space of permissible biomarker profiles to use as inclusion criteria in clinical trials. Such biomarker-based synchronization of patient’s disease stage would enable testing a drug in a more homogeneous population. This would in turn greatly increase the power of clinical trials in AD where it is common to see extreme levels of variability in patient trajectories [50,51].

## Data Availability

Deidentified data from the Alzheimer's Disease Neuroimaging Study is available to the general scientific community.

http://adni.loni.usc.edu/data-samples/access-data/

## Acknowledgement

Data collection and sharing for this project was funded by the Alzheimer’s Disease Neuroimaging Initiative (ADNI) (National Institutes of Health Grant U01 AG024904) and DOD ADNI (Department of Defense award number W81XWH-12-2-0012). ADNI is funded by the National Institute on Aging, the National Institute of Biomedical Imaging and Bioengineering, and through generous contributions from the following: AbbVie, Alzheimer’s Association; Alzheimer’s Drug Discovery Foundation; Araclon Biotech; BioClinica, Inc.; Biogen; Bristol-Myers Squibb Company; CereSpir, Inc.; Cogstate; Eisai Inc.; Elan Pharmaceuticals, Inc.; Eli Lilly and Company; EuroImmun; F. Hoffmann-La Roche Ltd and its affiliated company Genentech, Inc.; Fujirebio; GE Healthcare; IXICO Ltd.; Janssen Alzheimer Immunotherapy Research & Development, LLC.; Johnson & Johnson Pharmaceutical Research & Development LLC.; Lumosity; Lundbeck; Merck & Co., Inc.; Meso Scale Diagnostics, LLC.; NeuroRx Research; Neurotrack Technologies; Novartis Pharmaceuticals Corporation; Pfizer Inc.; Piramal Imaging; Servier; Takeda Pharmaceutical Company; and Transition Therapeutics. The Canadian Institutes of Health Research is providing funds to support ADNI clinical sites in Canada. Private sector contributions are facilitated by the Foundation for the National Institutes of Health (www.fnih.org). The grantee organization is the Northern California Institute for Research and Education, and the study is coordinated by them Alzheimer’s Therapeutic Research Institute at the University of Southern California. ADNI data are disseminated by the Laboratory for Neuro Imaging at the University of Southern California.

## Supplementary material

### Cholinesterase inhibitor treatment data extraction and processing

Treatment status with cholinesterase inhibitors was extracted from data on key background medications and the concurrent medications log. The latter was searched for the following common names and misspellings *‘aricept’, ‘aricpet’, ‘donepezil’, ‘exelon’, ‘galantamine’, ‘razadyne’, ‘remenyl’, ‘reminyl’, ‘rivastigmine’*.

Missing month and day in dates were interpolated as follows: missing day was set to the 1^st^ day of the month. Missing month was set to July.

**Supplementary Table 1.** Sex differences.

|  | Female | Male | p-value* |
| --- | --- | --- | --- |
| *Participants, n* | 1,002 | 1,140 | — |
| *ADAS-cog scores, n* | 4,364 | 5,466 | — |
| *Follow-up time (months),*<br>*median [IQR]* | 24.3 [5.5, 59.5] | 25.4 [11.9, 60.6] | 0.0085 |
| *Age at baseline (years),*<br>*median [IQR]* | 72.0 [57.4, 77.5] | 74.3 [69.5, 79.3] | < 0.0001 |
| *Length of education (years),*<br>*median [IQR]* | 15 [14, 18] | 16 [15, 18] | < 0.0001 |

\*p-values computed using Wilcoxon rank sum tests
IQR: Interquartile range

**Supplementary Table 2.** Parameter estimates in model adjusting for age, sex and education. p-values are computed using likelihood ratio tests.

| Parameter | Estimate | p-value |
| --- | --- | --- |
| *l* (intercept) | 0.01337 | — |
| *s* (significant memory concern) | -35.11 | 0.0106 |
| *s* (early MCI) | -44.58 | < 0.0001 |
| *s* (late MCI) | -87.73 | < 0.0001 |
| *s* (dementia) | -136.1 | < 0.0001 |
| *s* (age at baseline) | -1.878 | < 0.0001 |
| *s* (male) | 57.15 | < 0.0001 |
| *g* (intercept) | 3.169 | — |
| *g* (age at baseline) | 0.007061 | < 0.0001 |
| *g* (male) | -0.1895 | < 0.0001 |
| *g* (years of education) | -0.003337 | 0.0011 |
| *v* (intercept) | 7.623 | — |
| *v* (age at baseline) | 0.1234 | < 0.0001 |
| *v* (male) | 3.113 | < 0.0001 |
| *v* (years of education) | -0.4285 | < 0.0001 |

**Supplementary Table 3.** Parameter estimates in model adjusting for age, sex, education and biomarkers. p-values are computed using likelihood ratio tests.

| Parameter | Estimate | p-value |
| --- | --- | --- |
| *l* (intercept) | 17.92 | — |
| *s* (significant memory concern) | -18.20 | 0.0663 |
| *s* (early MCI) | -13.14 | < 0.0001 |
| *s* (late MCI) | -40.00 | < 0.0001 |
| *s* (dementia) | -64.12 | < 0.0001 |
| *s* (FDG-PET) | 73.52 | < 0.0001 |
| *s* (hippocampal volume) | 0.005980 | < 0.0001 |
| *s* (Florbetapir SUVR) | -34.57 | < 0.0001 |
| *s* (CSF Aβ1-42/Aβ1-40) | 137.1 | < 0.0001 |
| *s* (plasma NfL) | -0.2579 | < 0.0001 |
| *g* (intercept) | 3.280 | — |
| *g* (age at baseline) | 0.01276 | < 0.0001 |
| *g* (male) | -0.3764 | < 0.0001 |
| *g* (years of education) | -0.03897 | 0.0008 |
| *v* (intercept) | 11.71 | — |
| *v* (male) | 4.026 | < 0.0001 |
| *v* (years of education) | -0.2133 | 0.217 |

## References

1. Anderson RM, Hadjichrysanthou C, Evans S, Wong MM. Why do so many clinical trials of therapies for Alzheimer’s disease fail? The Lancet. 2017 Nov; 390(10110): 2327–2329.

2. Insel P, Weiner M, Mackin R, Mormino E, Lim Y, Stomrud E, et al. Determining clinically meaningful decline in preclinical Alzheimer disease. Neurology. 2019 Jul; 93(4): e322–e333.

3. Bateman R, Aisen P, De Strooper B, Fox N, Lemere C, Ringman J, et al. Autosomal-dominant Alzheimer’s disease: a review and proposal for the prevention of Alzheimer’s disease. Alzheimer’s research & therapy. 2011; 3(1): 1–13.

4. Ryman D, Acosta-Baena N, Aisen P, Bird T, Danek A, Fox N, et al. Symptom onset in autosomal dominant Alzheimer disease: a systematic review and meta-analysis. Neurology. 2014 Jul; 83(3): 253–260.

5. Wang G, Coble D, McDade E, Hassenstab J, Fagan A, Benzinger T, et al. Dominantly Inherited Alzheimer Network. Staging biomarkers in preclinical autosomal dominant Alzheimer’s disease by estimated years to symptom onset. Alzheimer’s & Dementia. 2019 Apr; 15(4): 506–514.

6. Bateman R, Benzinger T, Berry S, Clifford D, Duggan C, Fagan A, et al. The DIAN-TU Next Generation Alzheimer’s prevention trial: adaptive design and disease progression model. Alzheimer’s & Dementia. 2017 Jan; 13(1): 8–19.

7. Jack Jr C, Bennett D, Blennow K, Carrillo M, Dunn B, Haeberlein S, et al. NIA-AA Research Framework: Toward a biological definition of Alzheimer’s disease. Alzheimer’s & Dementia. 2018 Apr; 14(4): 535–562.

8. Ito K, Corrigan B, Zhao Q, French J, Miller R, Soares H, et al. Disease progression model for cognitive deterioration from Alzheimer’s Disease Neuroimaging Initiative database. Alzheimer’s & Dementia. 2011 Mar; 7(2): 151–160.

9. Gomeni R, Simeoni M, Zvartau-Hind M, Irizarry M, Austin D, Gold M. Modeling Alzheimer’s disease progression using the disease system analysis approach. Alzheimer’s & Dementia. 2012 Jan; 8(1): 39–50.

10. Samtani M, Farnum M, Lobanov V, Yang E, Raghavan N, DiBernardo A, et al. An improved model for disease progression in patients from the Alzheimer’s disease neuroimaging initiative. The Journal of Clinical Pharmacology. 2012 May; 52(5): 629–644.

11. Delor I, Charoin J, Gieschke R, Retout S, Jacqmin P, Alzheimer’s Disease Neuroimaging Initiative. Modeling Alzheimer’s disease progression using disease onset time and disease trajectory concepts applied to CDR-SoB scores from ADNI. CPT: pharmacometrics & systems pharmacology. 2013 Oct; 2(10): 1–10.

12. Donohue M, Jacqmin-Gadda H, Le Goff M, Thomas R, Raman R, Gamst A, et al. Estimating long-term multivariate progression from short-term data. Alzheimer’s & Dementia. 2014 Oct; 10(5): 200–410.

13. Li D, Iddi S, Thompson W, Rafii M, Aisen P, Donohue M. Alzheimer’s Disease Neuroimaging Initiative. Bayesian latent time joint mixed-effects model of progression in the Alzheimer’s Disease Neuroimaging Initiative. Alzheimer’s & Dementia: Diagnosis, Assessment & Disease Monitoring. 2018 Jan; 10: 657–668.

14. Raket L, Sommer S, Markussen B. A nonlinear mixed-effects model for simultaneous smoothing and registration of functional data. Pattern Recognition Letters. 2014 Mar; 38: 1–7.

15. Mohs R, Knopman D, Petersen R, Ferris S, Ernesto C, Grundman M, et al. Development of cognitive instruments for use in clinical trials of antidementia drugs: additions to the Alzheimer’s Disease Assessment Scale that broaden its scope. Alzheimer Disease & Associated Disorders. 1997; 11: 13/21.

16. Hughes C, Berg L, Danziger W, Coben L, Martin R. A new clinical scale for the staging of dementia. The British journal of psychiatry. 1982 Jun; 140(6): 566–572.

17. Pfeffer R, Kurosaki T, Harrah Jr C, Chance J, Filos S. Measurement of functional activities in older adults in the community. Journal of gerontology. 1982 May; 37(3): 323–329.

18. Landau S, Harvey D, Madison C, Koeppe R, Reiman E, Foster N, et al. Alzheimer’s Disease Neuroimaging Initiative. Associations between cognitive, functional, and FDG-PET measures of decline in AD and MCI. Neurobiology of aging. 2011 Jul; 32(7): 1207–1218.

19. Fischl B. FreeSurfer. Neuroimage. 2012 Aug; 62(2): 774–781.

20. Landau S, Fero A, Baker S, Koeppe R, Mintun M, Chen K, et al. Measurement of longitudinal β-amyloid change with 18F-florbetapir PET and standardized uptake value ratios. Journal of Nuclear Medicine. 2015 Apr; 56(4): 567–574.

21. Bittner T, Zetterberg H, Teunissen C, Ostlund Jr R, Militello M, Andreasson U, et al. Technical performance of a novel, fully automated electrochemiluminescence immunoassay for the quantitation of β-amyloid (1–42) in human cerebrospinal fluid. Alzheimer’s & Dementia. 2016 May; 12(5): 517–526.

22. Mattsson N, Cullen N, Andreasson U, Zetterberg H, Blennow K. Association between longitudinal plasma neurofilament light and neurodegeneration in patients with Alzheimer disease. JAMA neurology. 2019 Apr; 201976(7): 791–799.

23. Yang E, Farnum M, Lobanov V, Schultz T, Raghavan N, Samtani M, et al. Quantifying the pathophysiological timeline of Alzheimer’s disease. Journal of Alzheimer’s Disease. 2011 Jan; 26(4): 745–753.

24. Ramsay JO. Monotone regression splines in action. Statistical science. 1988; 3(4): 425–441.

25. Akaike H. Information theory and an extension of the maximum likelihood principle. In Selected papers of Hirotugu Aakaike. New York, NY: Springer; 1998. p. 199–213.

26. Schwarz G. Estimating the dimension of a model. The annals of statistics. 1978; 6(2): 461–464.

27. R Core Team. R: A language and environment for statistical computing. 2015..

28. Lindstrom M, Bates D. Nonlinear mixed effects models for repeated measures data. Biometrics. 1990 Sep;: 673–687.

29. Pinheiro J, Bates D, DebRoy S, Sarkar D, R Core Team. nlme: Linear and nonlinear mixed effects models. R package version 3.1-141. 2019..

30. Palmqvist S, Zetterberg H, Mattsson N, Johansson P, Minthon L, Blennow K, et al. Detailed comparison of amyloid PET and CSF biomarkers for identifying early Alzheimer disease. Neurology. 2015 Oct; 85(14): 1240–1249.

31. Embretson S, Reise S. Item response theory: Psychology Press; 2013.

32. Balsis S, Unger A, Benge J, Geraci L, Doody R. Gaining precision on the Alzheimer’s Disease Assessment Scale-cognitive: a comparison of item response theory-based scores and total scores. Alzheimer’s & Dementia. 2012 Jul; 8(4): 288–294.

33. Gardner R, Valcour V, Yaffe K. Dementia in the oldest old: a multi-factorial and growing public health issue. Alzheimer’s research & therapy. 2013 Aug; 5(4).

34. Stanley K, Whitfield T, Kuchenbaecker K, Sanders O, Stevens T, Walker Z. Rate of Cognitive Decline in Alzheimer’s Disease Stratified by Age. Journal of Alzheimer’s Disease. 2019 Jun; 69(4): 1153–1160.

35. Alzheimer’s Association. 2018 Alzheimer’s disease facts and figures. Alzheimer’s & Dementia. 2018 Mar; 14(3): 367–429.

36. Ferretti M, Iulita M, Cavedo E, Chiesa P, Dimech A, Chadha A, et al. Sex differences in Alzheimer disease—the gateway to precision medicine. Nature Reviews Neurology. 2018 Jul; 14(8): 457–469.

37. Oveisgharan S, Arvanitakis Z, Yu L, Farfel J, Schneider J, Bennett D. Sex differences in Alzheimer’s disease and common neuropathologies of aging. Acta neuropathologica. 2018 Dec; 136(6): 887–900.

38. Tucker A, Stern Y. Cognitive reserve in aging. Current Alzheimer Research. 2011 Jun; 8(4): 354–360.

39. Teri L, McCurry S, Edland S, Kukull W, Larson E. Cognitive decline in Alzheimer’s disease: a longitudinal investigation of risk factors for accelerated decline. The Journals of Gerontology Series A: Biological Sciences and Medical Sciences. 1995 Jan; 50(1): M49–M55.

40. Rasmusson D, Carson K, Brookmeyer R, Kawas C, Brandt J. Predicting rate of cognitive decline in probable Alzheimer’s disease. Brain and Cognition. 1996 Jun; 31(2): 133–147.

41. Wilson R, Li Y, Aggarwal N, Barnes L, McCann J, Gilley D, et al. Education and the course of cognitive decline in Alzheimer disease. Neurology. 2004 Oct; 63(7): 1198–1202.

42. Andel R, Vigen C, Mack W, Clark L, Gatz M. The effect of education and occupational complexity on rate of cognitive decline in Alzheimer’s patients. Journal of the International Neuropsychological Society. 2006 Jan; 12(1): 147–152.

43. Scarmeas N, Albert S, Manly J, Stern Y. Education and rates of cognitive decline in incident Alzheimer’s disease. Journal of Neurology, Neurosurgery & Psychiatry. 2006 Mar; 77(3): 308–316.

44. Musicco M, Palmer K, Salamone G, Lupo F, Perri R, Mosti S, et al. Predictors of progression of cognitive decline in Alzheimer’s disease: the role of vascular and sociodemographic factors. Journal of neurology. 2009 Aug; 256(8): 1288–1295.

45. Thomas R, Albert M, Petersen R, Aisen P. Longitudinal decline in mild-to-moderate Alzheimer’s disease: Analyses of placebo data from clinical trials. Alzheimer’s & Dementia. 2016 May; 12(5): 598–603.

46. Stern Y. Cognitive reserve in ageing and Alzheimer’s disease. The Lancet Neurology. 2012 Nov; 11(11): 1006–1012.

47. Birks J. Cholinesterase inhibitors for Alzheimer’s disease. Cochrane database of systematic reviews. 2006;(2).

48. Schneider L. Could cholinesterase inhibitors be harmful over the long term? International psychogeriatrics. 2012 Feb; 24(2): 171–174.

49. Kennedy R, Cutter G, Me F, Schneider L. Association of concomitant use of cholinesterase inhibitors or memantine with cognitive decline in alzheimer clinical trials: A meta-analysis. JAMA network open. 2018 Nov; 1(7): e184080.

50. Cummings J, Atri A, Ballard C, Boneva N, Frölich L, Molinuevo J, et al. Insights into globalization: comparison of patient characteristics and disease progression among geographic regions in a multinational Alzheimer’s disease clinical program. Alzheimer’s research & therapy. 2018 Dec; 10(1): 116.

51. Ballard C, Atri A, Boneva N, Cummings J, Frölich L, Molinuevo J, et al. Enrichment factors for clinical trials in mild-to-moderate Alzheimer’s disease. Alzheimer’s & Dementia: Translational Research & Clinical Interventions. 2019 Jan; 5: 164–174.

